# A User-Centered, SMART-on-FHIR-Based Dashboard Prototype for Diabetic Macular Edema

**DOI:** 10.64898/2026.09.15.26362892

**Authors:** Katja Hoffmann, Sophia Grummt, Hamed Mishian, Maximilian Nussbaumer, Markus Wolfien, Gabriel Stolze, Katrin Engelmann, Martin Sedlmayr

## Abstract

**Introduction:** Diabetic macular edema (DME) management requires integration of ophthalmic and systemic data, yet clinical decision-making remains limited by fragmented information systems and insufficient interoperability. This study aims to design a user-centered, interoperable dashboard prototype to support integrated clinical decision-making in DME care.

**Methods:** The prototype was developed using an agile, user-centered approach and implemented within a SMART-on-FHIR–compatible layered architecture. FHIR-based data standardization, including ophthalmology-specific profiling, enabled interoperable integration and visualization of structured clinical and imaging data within a modular web-based dashboard prototype.

**Results:** The resulting prototype provides a patient-centered dashboard integrating ophthalmic imaging findings and systemic risk factors, such as long-term glycemic markers, inflammatory markers, and blood pressure, within a unified interface. Built on standardized FHIR resources and SMART-on-FHIR authorization, the system demonstrates how user-centered dashboards can be implemented within interoperable, vendor-neutral healthcare IT infrastructures.

**Conclusion:** The presented design case illustrates how user-centered interface design and standards-based interoperability can jointly support the translation of clinical decision support tools into routine care environments. Establishing such interoperable infrastructures represents a key prerequisite for the sustainable deployment of future model-based clinical decision support systems.

## 1. Introduction

Diabetic macular edema (DME) is a leading cause of vision impairment among adults with diabetes mellitus [1]. Its management requires individualized decisions based on retinal imaging findings as well as systemic metabolic parameters, such as glycated hemoglobin (HbA1c), inflammatory markers (e.g., hs-CRP), and blood pressure [2]. Effective treatment therefore depends on interdisciplinary collaboration and the integration of ophthalmic and systemic patient information, reflecting the health of the entire body [3].

In routine practice, however, clinical decision-making often remains focused on ocular findings, while relevant systemic biomarkers, such as HbA1c, are stored in separate information systems and are not readily accessible. This fragmentation contrasts with the multifactorial nature of DME and limits holistic care. Clinical decision support systems (CDSS) can help bridge this gap by integrating data into a joint platform, resulting in information that are user-friendly, intuitive, and seamlessly integrated into clinical workflows [4].

Designing interoperable CDSS extends beyond interface development and requires a vendor-neutral, standards-based infrastructure capable of aggregating heterogeneous clinical data. At the same time, User-Centered Design (UCD) principles [5] are essential to ensure usability and routine adoption. Integrated dashboards must therefore combine robust technical integration with intuitive, workflow-oriented user interfaces.

The dashboard presented here serves as a CDSS design case for such an interoperable infrastructure. It integrates ophthalmic and systemic findings within a vendor-neutral architecture to enable structured cross-system data exchange. The primary contribution lies in demonstrating how interoperable infrastructure can support decision-making in DME care, focusing on architectural and integration aspects rather than clinical outcome evaluation while remaining applicable beyond this specific clinical use case. The guiding research question is:

*How can an interoperable, standards-based clinical dashboard be designed and implemented as part of a vendor-neutral infrastructure to integrate ophthalmic and systemic biomarkers (e.g., HbA1c, hs-CRP, and blood pressure) for DME care?*

## 2. Methods

### 2.1. Agile Development and Technical Framework

The DME Dashboard prototype was developed using an agile, iterative methodology enabling incremental refinement from a *minimal viable product (MVP)* towards extended versions, following the approach described in our previous work [6]. This study focuses on the *initial MVP*, which establishes the foundational architecture for an interoperable dashboard.

The system architecture follows a layered SMART-on-FHIR compatible design separating four core components: (i) the FHIR server BLAZE [7] providing structured patient data via HL7 FHIR resources and profiles (see Section 2.3), (ii) the identity and access management system Keycloak v26.4.7 [8] handling authentication and authorization via OAuth 2.0 [9] and OpenID Connect [10], (iii) the web-based dashboard prototype, and (iv) the Proxy SMART middleware v0.0.2-alpha [11], which enables SMART-on-FHIR authorization flows by acting as an intermediary between the dashboard application and the backend.

Development of the dashboard prototype proceeded through the sprint-based phases comprising requirement analysis and detailed software design (*Design*, see Section 2.2), data standardization based on HL7 FHIR (*Unify*, see Section 2.3), and implementation of the software units (*Build*, see Section 2.4). Figure 1 provides a visual overview of these development phases alongside the architectural framework.

**Figure 1.**
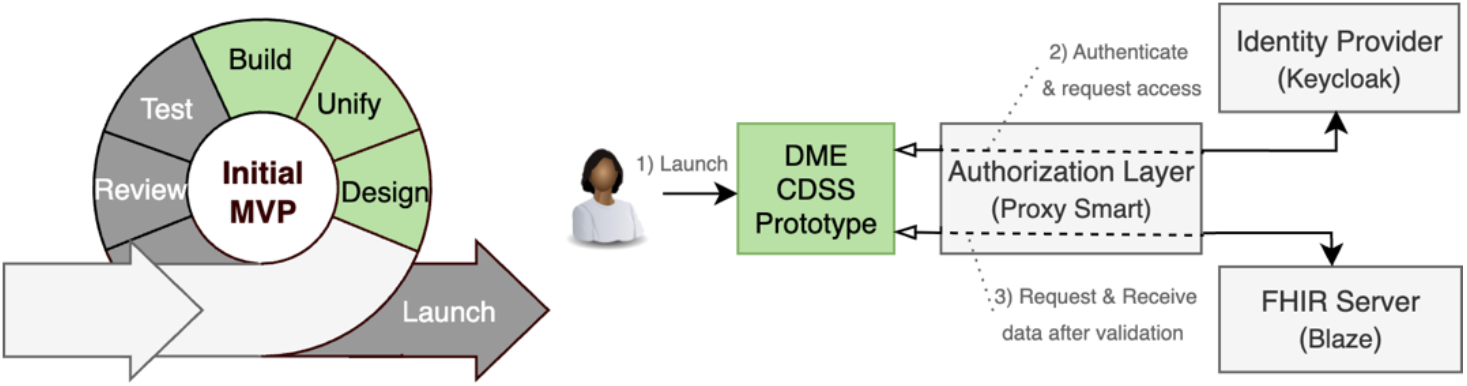
Development phases and technical framework of the Diabetic Macular Edema (DME) dashboard

### 2.2. User-Centered Design

A structured UCD approach was applied to derive clinical and workflow requirements. The overall design process, including structured discussions, interviews, persona development, low- and mid-fidelity mockups, and iterative feedback cycles with ophthalmologists, has been described in detail in Grummt et al. [12]. Building on this foundation, the design concepts were translated into a functional dashboard prototype, while iterative feedback from clinicians continued during implementation through four refinement cycles. The design aimed to visually represent parameter development and disease progression over time to support assessment of treatment effectiveness. Physicians also requested that missing data be highlighted to provide a complete view of patient information and that DME-relevant non-ophthalmological parameters, such as HbA1c and blood pressure, be included to support multidisciplinary assessment. This iterative UCD process informed both the structural layout of the dashboard and the prioritization of displayed parameters.

### 2.3. Unification through Data Standardization and FHIR Profiling

FHIR-based data modeling was guided by the German Medical Informatics Initiative (MII) core data set (CDS) [13]. While existing profiles sufficiently represent general entities (e.g., Patient, Condition, Procedure), ophthalmology-specific parameters are not sufficiently covered [14]. To address this gap, ophthalmic-specific FHIR profiles developed within the MII project *EyeMatics* were adopted [15]. For parameters not yet covered, project-specific FHIR specifications were defined to enable structured and interoperable representation within the prototype [16].

### 2.4. Technical Implementation of the DME Dashboard in the Build Phase

The functional DME Dashboard prototype was implemented as a web-based application using the Next.js framework v14.2.35 [17], enabling modular and scalable frontend development. The application dynamically retrieves and processes structured HL7 FHIR resources according to the specifications described in Section 2.3.

The dashboard prototype is implemented as a SMART-on-FHIR provider application, allowing clinicians to launch the DME Dashboard. To demonstrate the Dashboard prototype independently of an Electronic Health Record (EHR) system, the standalone approach was chosen for the prototype implementation, but it can be adapted to an EHR-integrated launch context at any time. This launch mode follows the SMART App Launch specification for *Provider Apps that Launch Standalone* as defined by the SMART-on-FHIR implementation guide v2.2 [18].

## 3. Results

The implemented DME Dashboard provides a consolidated, patient-centered overview of clinically relevant parameters for disease monitoring (Figure 2).

**Figure 2.**
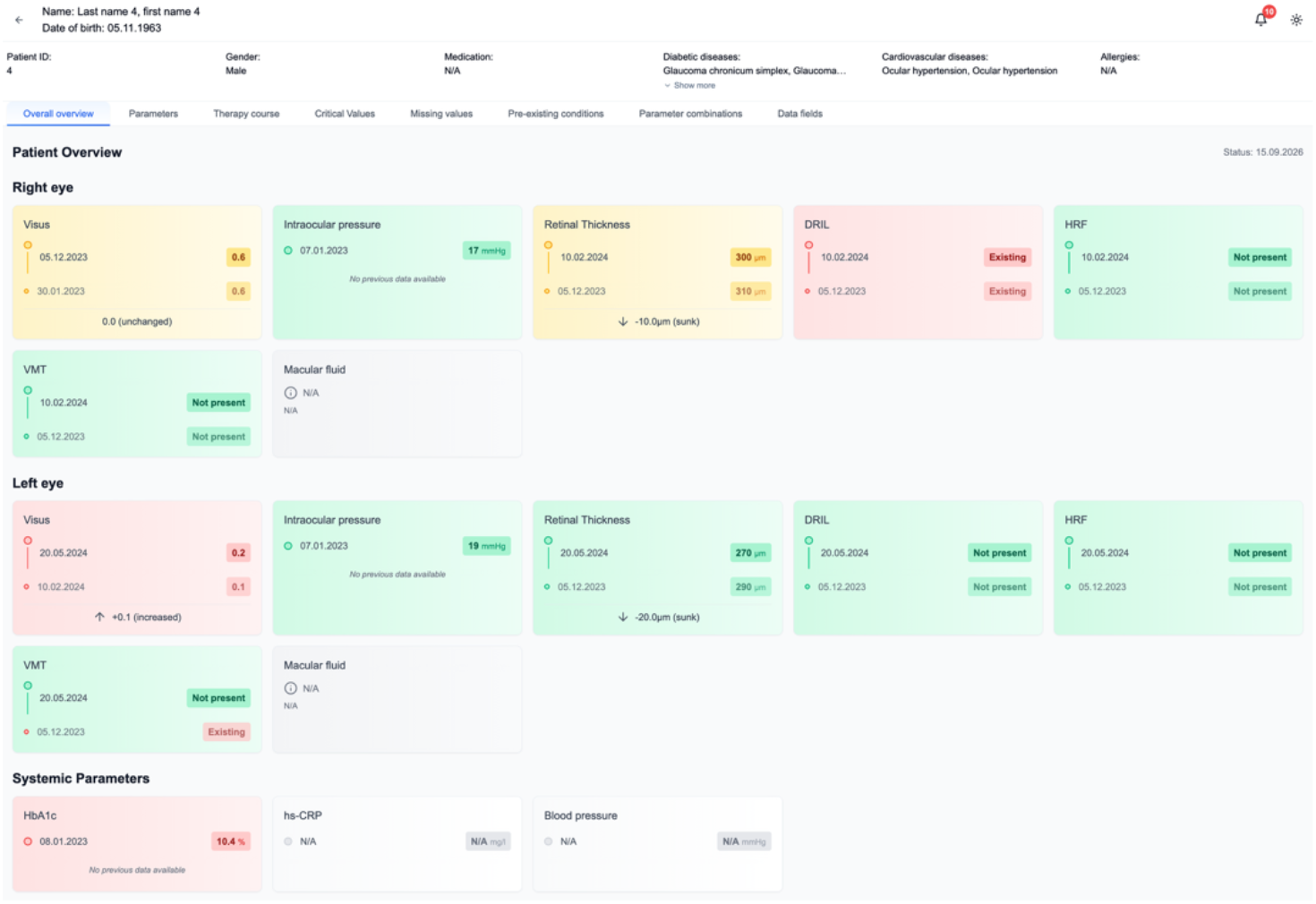
Patient-level overview of the implemented DME dashboard prototype showing the most recent and previous ophthalmic and systemic measurements with trend indicators and color-coded clinical thresholds (grey:missing; green:normal; yellow:borderline; red:outside normal range).

The interface aggregates visual acuity, intraocular pressure, retinal OCT-derived biomarkers (e.g., retinal thickness, hyperreflective foci, and other morphological indicators), as well as the relevant systemic risk factors HbA1c, hs-CRP, and blood pressure. For each parameter, the most recent and previous measurements are displayed side-by-side together with the calculated difference, enabling rapid comparison between visits. Trend indicators and color-coded thresholds (green, yellow, red) highlight clinically relevant deviations and support rapid interpretation of disease status.

In addition to the parameter overview, the dashboard provides a longitudinal visualization of treatment history and clinical outcomes (Figure 3). Visual acuity measurements are displayed along a temporal axis together with documented therapeutic interventions, such as intravitreal injections or other treatment modalities. This integrated representation allows clinicians to relate functional outcomes to previous treatment decisions and facilitates recognition of response patterns over time.

**Figure 3.**
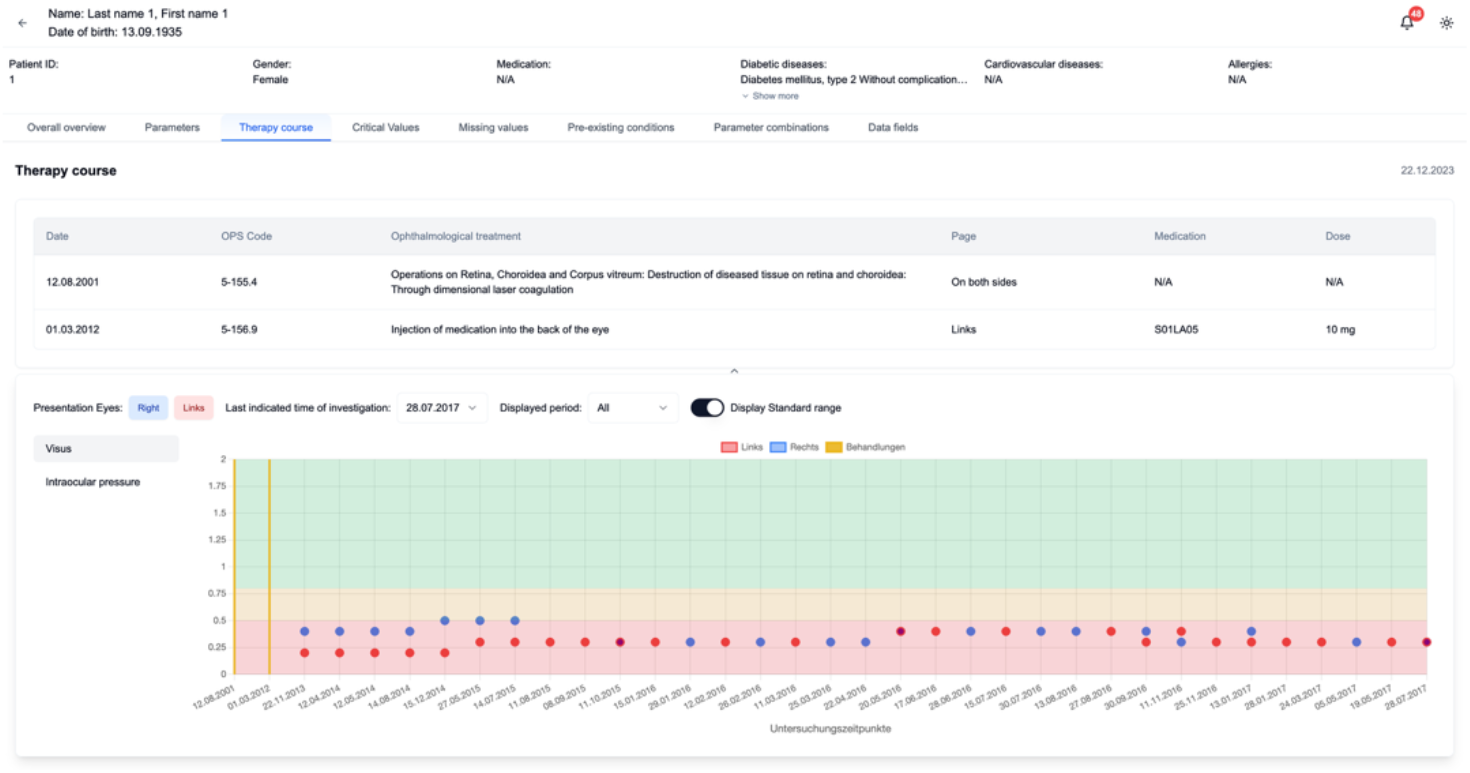
Longitudinal visualization of visual acuity measurements in relation to documented therapeutic interventions of the implemented DME dashboard prototype.

Missing or non-documented parameters are explicitly marked within the interface, enabling transparent assessment of data completeness and preventing implicit assumptions regarding the availability of clinical information.

In addition to the dashboard interface, the implemented prototype includes a SMART-on-FHIR–compatible demonstrator infrastructure supporting interoperable access to clinical data. The system integrates a FHIR server (Blaze), an identity and access management system (Keycloak), and the Proxy SMART middleware implementing the SMART-on-FHIR authorization flow as described in Section 2.1.

To ensure reproducibility and experimentation, the complete prototype implementation, including deployment source code of the implemented dashboard prototype, deployment configuration, and synthetic test data, is provided as an open-source repository [19]. Complementary design artifacts from the UCD process, including personas and mid-fidelity mockups, are available as supplementary material via Zenodo (DOI: 10.5281/zenodo.19194608). The repository includes the dashboard application, infrastructure configuration files for deploying the system components, and a synthetic clinical dataset for testing the prototype.

## 4. Discussion

This work aimed to design a user-centered, interoperable dashboard to support clinical decision-making in DME. The presented prototype demonstrates how ophthalmic and systemic patient data can be integrated within a standards-based, vendor-neutral infrastructure to provide a consolidated view of clinically relevant parameters. Beyond the implementation of a functional dashboard interface, the project illustrates architectural and methodological considerations relevant for scalable and interoperable CDSS development.

First, the strict separation of data provisioning, authorization services, and application logic proved essential for maintaining modularity and extensibility. In alignment with emerging SMART-on-FHIR frameworks, this layered approach enables flexible integration into heterogeneous healthcare IT environments. In particular, the use of a Proxy SMART middleware allowed the realization of SMART-on-FHIR-compatible authorization flows even in infrastructures that do not natively support SMART-on-FHIR. By mediating between the application, FHIR server, and identity provider, this approach enables a vendor-neutral integration strategy and lowers adoption barriers for existing clinical systems, as previously described in our work on vendor-neutral SMART-on-FHIR infrastructures [20]. Such architectural decoupling may therefore represent an important prerequisite for more sustainable dashboard and CDSS deployment across institutional boundaries.

Second, the project highlights that data standardization is not merely a technical requirement but a central design driver. While existing MII CDS [13] and *EyeMatics* profiles [15] provided a valuable foundation for representing clinical entities, domain-specific extensions were necessary to adequately capture ophthalmological biomarkers. This observation underlines the importance of iterative profile development and active contribution to evolving interoperability standardization initiatives when implementing specialized clinical applications.

Third, the use of an agile MVP strategy enabled early validation of architectural decisions while reducing implementation risks. By establishing the technical framework prior to extensive feature expansion, the project prioritized structural robustness over premature functional complexity. This approach may facilitate the gradual introduction of interoperable CDSS solutions within complex clinical IT ecosystems.

Although the present work focuses on the implementation of a clinical dashboard, the underlying infrastructure is intended to support the integration of computational decision-support models in future system expansions. As outlined in our previous work on a technological concept for integrating AI-based CDSS into clinical workflows [6], the development strategy follows staged expansion levels ranging from an initial MVP to progressively more advanced decision-support functionalities. Within this framework, the interoperable infrastructure established in the present study represents the foundational stage enabling subsequent integration of computational models. These may include machine learning–based approaches as well as knowledge-based or statistical models, depending on the clinical application. Establishing a standards-based infrastructure at an early stage therefore represents a prerequisite for the sustainable integration of diverse model-based CDSS components.

Several limitations must be acknowledged. The presented prototype represents an MVP and has not yet undergone integration into routine clinical infrastructure. Formal evaluation of usability, workflow integration, and user acceptance remains part of future work. However, the dashboard was developed in close collaboration with ophthalmologists through an iterative UCD process, ensuring that clinical requirements and workflow considerations were incorporated throughout development.

While the current work focuses on ophthalmology, the underlying framework is not domain-restricted. The combination of FHIR-based data modeling, vendor-neutral SMART-on-FHIR infrastructure, and modular application design may therefore serve as a transferable blueprint for interoperable CDSS development in other medical domains.

## 5. Conclusion

This study presented a user-centered, interoperable dashboard designed to support ophthalmologists in clinical decision-making for DME. By integrating ophthalmic and systemic parameters within a SMART-on-FHIR–compatible architecture, the prototype demonstrates how clinically relevant data can be consolidated across heterogeneous systems in a vendor-neutral manner. The design case highlights the value of combining user-centered design with standards-based interoperability to advance scalable clinical decision-support infrastructures. Future research should focus on clinical deployment and evaluation in routine care. Overall, the presented design case provides practical insights into how interoperable infrastructures and user-centered design can jointly support the translation of clinical decision-support tools into real-world healthcare environments, illustrated through the development of an interoperable dashboard for DME care.

## Data Availability

All data and materials referred to in this manuscript are openly available. The complete prototype implementation, including the dashboard application, deployment configuration, and synthetic test data, is available as an open-source repository at https://gitlab.ukdd.de/pub/mihubx/smart-on-fhir-ophthalmic-stack. Complementary design artifacts from the user-centered design process, including personas and mid-fidelity mockups, are available as supplementary material via Zenodo (DOI: 10.5281/zenodo.19194608).

## Declarations

### Author contributions

KH: Conceptualization, Visualization, Writing – original draft. KH, SG: Methodology, Investigation, Formal Analysis. HM, KH: Software. MN, GS, KE: Resources. GS, KE: Validation (Clinical). MS: Supervision. All authors: Writing – review & editing. All authors approved the manuscript in the submitted version and take responsibility for the scientific integrity of the work.

## Acknowledgement

This work is part of the projects “Medical Informatics Hub in Saxony (MiHUBx)” (Grant Numbers 01ZZ2101A and 01ZZ2101E), funded by the German Federal Ministry of Research and Technology (BMFTR), and “Medical Informatics Hub (MiHUB)” (Grant Number 01ZZ2506A), funded by the German Ministry of Research, Technology and Space.

